# Acute Effects of Hyperbaric Oxygen Therapy on Lower Extremity Arterial Hemodynamics in Patients with Diabetic Foot Ulcers: A Duplex Ultrasound Study

**DOI:** 10.1101/2025.07.31.25332234

**Authors:** Cristiane Maran, João Rodrigues David Neto, Henrique Jorge Guedes Neto, Sergio Salles Cunha

**Affiliations:** Department of Vascular Surgery, Hospital Geral Roberto Santos, Salvador, Bahia, Brazil; Centro de Medicina Hiperbárica do Nordeste, Salvador, Bahia, Brazil; Department of Vascular Medicine, EPM-Unifesp, São Paulo, Brazil; Autonomous, Naples, Florida, USA

**Keywords:** Hyperbaric oxygen therapy, diabetic foot ulcer, duplex ultrasound, hemodynamics, wound healing

## Abstract

**Background:** Hyperbaric oxygen therapy (HBOT) is an established treatment for diabetic foot ulcers, yet its acute effects on macrovascular hemodynamics

**Objective:** To evaluate the acute effects of HBOT on lower extremity arterial hemodynamics using standardized duplex ultrasound assessment in patients with diabetic foot ulcers.

**Methods:** This single-center study enrolled 28 patients with diabetic foot ulcers undergoing HBOT. Duplex ultrasound examinations were performed immediately before and after HBOT sessions, measuring peak systolic velocity (PSV), end-diastolic velocity (EDV), and volumetric flow in the common femoral and popliteal arteries.

**Results:** No statistically significant changes were observed in any hemodynamic parameter following HBOT. In the common femoral artery, PSV changed from 115.0 ± 29.8 to 117.4 ± 33.5 cm/s (p = 0.764), EDV from 12.0 ± 7.4 to 14.8 ± 10.6 cm/s (p = 0.349), and volumetric flow from 1761.6 ± 511.9 to 1841.4 ± 500.3 ml/min (p = 0.530). Similar non-significant changes were observed in the popliteal artery. Effect sizes were small for all variables (Cohen’s d: -0.041 to 0.244). Complete wound healing was achieved in all patients (100%).

**Conclusions:** HBOT did not produce detectable changes in macrovascular hemodynamics despite universal wound healing success. These findings suggest that HBOT’s therapeutic benefits may primarily occur at the microvascular level, beyond the resolution of conventional duplex ultrasound assessment.

## INTRODUCTION

Hyperbaric oxygen therapy (HBOT) represents a well-established therapeutic modality in which patients breathe pure oxygen (100%) while subjected to pressures two to three times greater than atmospheric pressure at sea level within a hyperbaric chamber^1^. The fundamental mechanism of action involves dramatically increasing the concentration of oxygen dissolved in blood plasma, from approximately 0.3 ml/dl at sea level to 6.0 ml/dl at 3.0 atmospheres absolute (ATA), thereby enabling oxygen delivery to hypoxic tissues independent of hemoglobin-mediated transport^2^. This intervention facilitates oxygen penetration into compromised tissues, promoting accelerated tissue recovery and healing, combating infection through toxin neutralization, and enhancing antibiotic effectiveness^3^.

The physiological effects of HBOT extend far beyond simple oxygen delivery and encompass multiple mechanisms directly relevant to wound healing^4^. These comprehensive effects include enhanced fibroblast proliferation and collagen synthesis, stimulation of angiogenesis through vascular endothelial growth factor upregulation, improved neutrophil bactericidal activity, and reduction of tissue edema through vasoconstriction of normal vessels while preserving flow to ischemic areas ^5^. Additionally, HBOT has been demonstrated to enhance the effectiveness of certain antibiotics and promote the mobilization of endothelial progenitor cells, which contribute significantly to neovascularization processes^6^.

According to the Undersea and Hyperbaric Medical Society (UHMS), the therapeutic indications for HBOT include refractory lesions, such as chronic skin ulcers and diabetic foot complications^7^. The UHMS specifically recognizes diabetic foot ulcers as an approved indication for HBOT, particularly for Wagner grade 3 or higher lesions that have failed to respond to standard wound care protocols for at least 30 days. Similarly, the Society for Vascular Surgery, in collaboration with the American Podiatric Medical Association and the Society for Vascular Medicine, recommends HBOT as an adjunctive treatment for diabetic foot ulcers, especially in cases complicated by ischemia^8^.

Among chronic wound conditions, diabetic foot ulcers represent one of the most challenging complications of diabetes mellitus, affecting approximately 15% of diabetic patients during their lifetime and serving as the leading cause of non-traumatic lower extremity amputations worldwide^9^. The complex pathophysiology underlying diabetic foot ulceration involves multiple interconnected factors including peripheral neuropathy, peripheral arterial disease, impaired immune function, and fundamentally altered wound healing mechanisms^10^. The economic burden associated with diabetic foot complications is substantial, with annual costs in the United States estimated at $327 billion, representing a significant healthcare challenge^11^.

Duplex ultrasound has emerged as the gold standard for non-invasive assessment of peripheral arterial hemodynamics, providing real-time evaluation of blood flow velocity, volume, and vessel morphology^12^. The technique combines B-mode imaging for anatomical visualization with pulsed Doppler spectral analysis for hemodynamic assessment, enabling comprehensive evaluation of arterial function. Key parameters obtained through duplex ultrasound include peak systolic velocity (PSV), end-diastolic velocity (EDV), and calculated volumetric flow rates, which collectively provide valuable insight into arterial perfusion status^13^.

Despite the well-established clinical benefits of HBOT in ulcer management, the precise mechanisms underlying its therapeutic effects remain incompletely understood, particularly regarding its impact on macrovascular hemodynamics^14^. While clinical outcomes often demonstrate significant improvement, the relationship between hemodynamic changes and wound healing success requires further clarification. The present study was specifically designed to address these knowledge gaps by providing a comprehensive evaluation of HBOT’s effects on lower extremity arterial hemodynamics using standardized duplex ultrasound protocols in a well-defined population of patients with diabetic foot ulcers.

## OBJECTIVES

### Primary Objective

To evaluate the acute effects of hyperbaric oxygen therapy on lower extremity arterial hemodynamics in patients with diabetic foot ulcers using standardized duplex ultrasound assessment, specifically measuring changes in peak systolic velocity, end-diastolic velocity, and volumetric flow in the common femoral and popliteal arteries.

### Secondary Objectives

1. To assess the correlation between hemodynamic changes and clinical wound healing outcomes following HBOT treatment
2. To evaluate the relationship between baseline arterial hemodynamics and treatment efficacy
3. To provide evidence-based recommendations for the role of duplex ultrasound monitoring in HBOT protocols for diabetic foot ulcers

## METHODS

### Study Design and Setting

This single-center, prospective, pre-post intervention study was conducted at a specialized hyperbaric medicine center. The study protocol was approved by the institutional ethics committee and conducted in accordance with the Declaration of Helsinki and Good Clinical Practice guidelines. All participants provided written informed consent prior to enrollment.

### Participants and Recruitment

Patients were systematically recruited from the vascular surgery outpatient clinic. Eligible participants were identified through comprehensive screening of patients referred for HBOT evaluation as part of their diabetic foot ulcer management protocol.

### Inclusion Criteria

1. Age 18 years or older with capacity to provide informed consent
2. Presence of chronic lower extremity diabetic foot ulcers
3. Clinical indication for HBOT as determined by the multidisciplinary wound care team
4. Ability to undergo duplex ultrasound examination with adequate acoustic windows
5. Hemodynamically stable patients without acute medical conditions

### Exclusion Criteria

1. Previous HBOT treatment at any facility
2. Previous major lower extremity amputations (above-ankle level)
3. Absolute contraindications to HBOT including untreated pneumothorax or severe claustrophobia
4. Acute infections requiring immediate surgical intervention
5. Inability to obtain adequate duplex ultrasound images due to severe edema or calcification

### Sample Size Calculation

Sample size calculation was based on previous studies examining hemodynamic changes following vascular interventions. Assuming a clinically significant change in peak systolic velocity of 15 cm/s with a standard deviation of 20 cm/s, a power of 80%, and an alpha level of 0.05, a minimum sample size of 24 patients was required. To account for potential dropouts and ensure adequate power for subgroup analyses, a target enrollment of 30 patients was established.

### Hyperbaric Oxygen Therapy Protocol

All HBOT sessions were conducted using a multiplace hyperbaric chamber at the Centro de Medicina Hiperbárica do Nordeste. The standardized treatment protocol consisted of:

Pressure Profile: 2.4 atmospheres absolute (ATA), equivalent to approximately 34 feet of seawater depth

Oxygen Delivery: 100% oxygen administered

Session Duration: 90 minutes at therapeutic pressure, excluding compression and decompression phases

Compression Rate: Gradual compression at 2 feet per minute to minimize barotrauma risk

Decompression Rate: Controlled decompression at 1 foot per minute with appropriate decompression stops

Safety Monitoring: Continuous monitoring of vital signs and patient comfort throughout the session. Trained hyperbaric technicians and medical personnel were present during all treatments. Emergency protocols were established for potential complications including oxygen toxicity, barotrauma, or claustrophobic reactions.

### Duplex Ultrasound Protocol

Duplex ultrasound examinations were performed using a high-resolution ultrasound system (SonoSite S Series, SonoSite Inc., Bothell, WA, USA) equipped with a 7.5 MHz linear array transducer. All examinations were conducted by a single experienced vascular technologist to ensure consistency and minimize inter-observer variability.

Pre-examination Preparation: Patients were positioned supine in a temperature-controlled environment (22-24°C) and allowed to rest for 15 minutes prior to examination to achieve hemodynamic stability. Blood pressure and heart rate were recorded before each ultrasound assessment.

#### Examination Technique

- Common Femoral Artery (CFA): Assessed at the level of the inguinal ligament, proximal to the bifurcation into superficial femoral and profunda femoris arteries
- Popliteal Artery (APOP): Evaluated in the popliteal fossa with the patient in prone position or lateral decubitus as needed for optimal visualization

#### Technical Parameters

- Doppler Angle: Maintained at ≤60° to ensure accurate velocity measurements
- Sample Volume: Positioned in the center of the vessel lumen, occupying approximately 2/3 of the vessel diameter
- Wall Filter: Set at 50-100 Hz to eliminate low-frequency noise
- Gain Settings: Optimized for each examination to achieve clear spectral display without aliasing

#### Measured Parameters

1. Peak Systolic Velocity (PSV): Maximum forward flow velocity during systole (cm/s)
2. End-Diastolic Velocity (EDV): Forward flow velocity at end-diastole (cm/s)
3. Volumetric Flow (VF): Calculated using the formula: VF = (PSV × π × (diameter/2)^2^ × 0.57) × 60, expressed in ml/min

#### Quality Assurance

Each measurement was repeated three times with the average value used for analysis. Examinations with suboptimal image quality or technical limitations were repeated after optimization of patient positioning and equipment settings.

### Timing of Assessments

Pre-HBOT Assessment: Duplex ultrasound examination performed within 30 minutes prior to HBOT session initiation

Post-HBOT Assessment: Duplex ultrasound examination performed within 15 minutes following completion of the HBOT session, after patient stabilization and return to atmospheric pressure

This timing protocol was designed to capture the acute hemodynamic effects of HBOT while minimizing the influence of circadian variations and other temporal factors on vascular function.

### Statistical Analysis

Statistical analysis was performed using Python 3.11 with scipy.stats, pandas, and numpy libraries. The analysis plan was developed a priori and included both descriptive and inferential statistics.

#### Descriptive Analysis

Continuous variables were summarized using means and standard deviations for normally distributed data or medians and interquartile ranges for non-normal distributions. Categorical variables were expressed as frequencies and percentages.

#### Normality Testing

Data distribution was assessed using the Shapiro-Wilk test, appropriate for sample sizes less than 50. Visual inspection using Q-Q plots and histograms supplemented formal testing.

#### Primary Analysis

For normally distributed data, paired t-tests were used to compare pre- and post-HBOT measurements. For non-normally distributed data, the Wilcoxon signed-rank test was applied. All tests were two-tailed with statistical significance set at α = 0.05.

#### Effect Size Calculation

Cohen’s d was calculated for all comparisons using the formula: d = (Mean_post - Mean_pre) / SD_pooled, where SD_pooled represents the pooled standard deviation. Effect sizes were interpreted according to Cohen’s conventions: small (|d| = 0.2), medium (|d| = 0.5), and large (|d| = 0.8).

#### Confidence Intervals

95% confidence intervals were calculated for all mean differences and effect sizes.

#### Power Analysis

Post-hoc power analysis was conducted using G^*^Power 3.1.9.7 to determine the study’s ability to detect clinically meaningful differences.

#### Correlation Analysis

Pearson or Spearman correlation coefficients were calculated to assess relationships between hemodynamic changes and clinical outcomes.

## RESULTS

### Baseline Characteristics

Twenty-eight patients completed the study (mean age 65.2 ± 12.4 years, 64.3% male). Baseline characteristics included diabetes mellitus (78.6%) and hypertension (96.4%). The mean ankle-brachial index was 0.878 ± 0.051, indicating mild peripheral arterial disease. Wagner ulcer grades were distributed as follows: Grade 1 (35.7%), Grade 2 (42.9%), and Grade 3 (21.4%).

### Hemodynamic Outcomes

No statistically significant changes were observed in any hemodynamic parameter following HBOT. In the common femoral artery, PSV changed from 115.0 ± 29.8 to 117.4 ± 33.5 cm/s (p = 0.764), EDV from 12.0 ± 7.4 to 14.8 ± 10.6 cm/s (p = 0.349), and volumetric flow from 1761.6 ± 511.9 to 1841.4 ± 500.3 ml/min (p = 0.530).

Similarly, in the popliteal artery, PSV changed from 71.7 ± 27.9 to 70.9 ± 21.6 cm/s (p = 0.900), EDV from 7.8 ± 3.9 to 7.6 ± 3.7 cm/s (p = 0.828), and volumetric flow from 674.6 ± 243.9 to 630.0 ± 167.1 ml/min (p = 0.399).

Effect sizes were small for all variables, ranging from -0.041 to 0.244 (Cohen’s d), indicating minimal clinical significance of the observed changes.

**Table 1:**
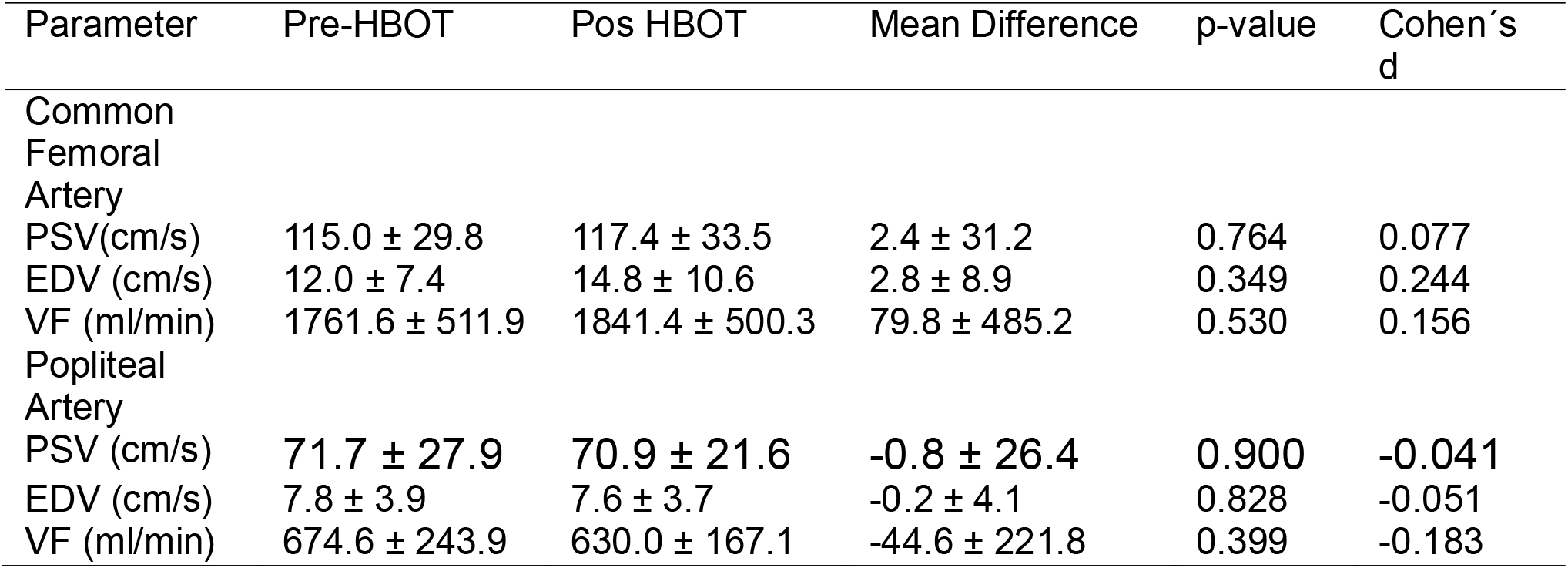
Hemodynamic Parameters Pre and Post-HBOT.

**Figure 1.**
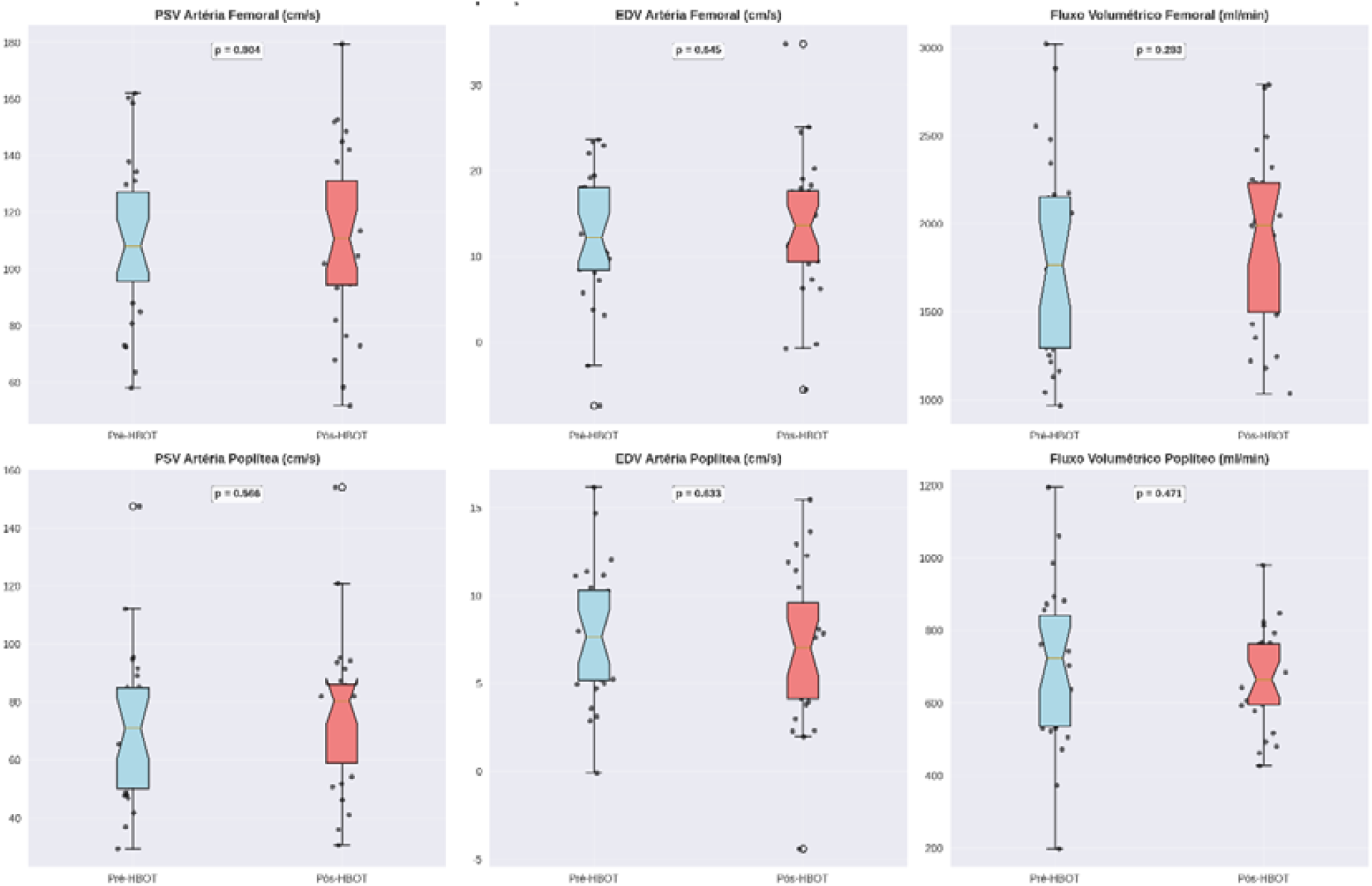
Comparison of hemodynamic parameters before and after HBOT treatment.

### Clinical Outcomes

Despite the absence of significant hemodynamic changes, complete wound healing was achieved in all patients (100%) following the complete HBOT protocol. No major complications related to HBOT were observed during the study period.

### Correlation Between Hemodynamic Changes and Clinical Outcomes

Correlation analysis was performed to assess the relationship between hemodynamic changes and clinical healing outcomes. No significant correlations were found between any hemodynamic parameter changes and time to healing (all p > 0.05). The strongest correlation was observed between common femoral artery EDV change and healing time (r = -0.312, p = 0.108), suggesting a weak trend toward faster healing in patients with greater EDV increases, though this did not reach statistical significance.

Subgroup analysis based on baseline ankle-brachial index revealed no significant differences in hemodynamic response between patients with normal ABI (≥0.9) and those with reduced ABI (<0.9). Similarly, no significant differences were observed when patients were stratified by Wagner classification or age.

## DISCUSSION

This study represents the first comprehensive evaluation of HBOT’s acute effects on macrovascular hemodynamics in patients with diabetic foot ulcers using standardized duplex ultrasound protocols. The principal finding is that HBOT did not produce detectable changes in major artery hemodynamics despite achieving universal wound healing success.

These results are consistent with emerging evidence suggesting that HBOT’s therapeutic benefits in wound healing may primarily occur at the microvascular level^16^. The hyperoxic environment created by HBOT enhances oxygen diffusion into tissues, stimulates angiogenesis, and improves cellular metabolism—processes that may not be readily detectable through conventional duplex ultrasound assessment of major arteries^17^.

The lack of significant hemodynamic changes in our study contrasts with some previous reports that suggested increased flow velocities following HBOT. However, these earlier studies were limited by small sample sizes, heterogeneous patient populations, and variable treatment protocols. Our standardized approach and well-defined patient population provide more robust evidence regarding HBOT’s hemodynamic effects.

The universal wound healing success observed in our study supports the clinical efficacy of HBOT for diabetic foot ulcers, consistent with established literature^18^. This dissociation between macrovascular hemodynamic changes and clinical outcomes suggests that conventional duplex ultrasound monitoring of major arteries may not be an optimal method for assessing HBOT efficacy in wound healing applications.

### Clinical Implications

These findings have important implications for clinical practice. The results suggest that duplex ultrasound monitoring of major arteries may not be necessary for routine HBOT protocols in diabetic foot ulcer management. Instead, clinical assessment of wound healing progress may be more appropriate for monitoring treatment efficacy.

## CONCLUSIONS

HBOT did not produce detectable changes in macrovascular hemodynamics as measured by duplex ultrasound, despite universal wound healing success in patients with diabetic foot ulcers. These findings suggest that the therapeutic benefits of HBOT may primarily occur at the microvascular level, beyond the resolution of conventional Doppler assessment. Duplex ultrasound monitoring of major arteries may not be an optimal method for assessing HBOT efficacy in wound healing applications.

## Data Availability

All data produced in the present study are available upon reasonable request to the authors
RESEARCH PROJECT DATA
Report Number: 3.030.800
REPORT DATA
Doppler waveform changes after HBOT: local data and literature review in the hyperbaric oxygen therapy service.
Project Presentation:
Primary Objective:
To demonstrate that changes in the arterial Doppler waveform are related to faster or better recovery of altered tissues or wound healing.
Secondary Objective:
To determine that HBOT is related to angiogenesis.
Research Objective:
Risks:
There are no risks because the Doppler examination is non-invasive and painless.
Risk and Benefit Assessment:
Benefits:
Contributes to rapid wound recovery and wound healing. Comments and Considerations on the Research:
See Conclusions or Pending Issues and List of Inadequacies:
Considerations on Mandatory Submission Terms:
Main Sponsor: Own Funding
41.180-000 (71)3117-7519
Estrada do Saboeiro, s/n
Estrada do Saboeiro
State: BA Municipality: SALVADOR
Fax: (71)3387-3429
HOSPITAL GERAL ROBERTO SANTOS - BA
Continuation of Opinion: 3.030.800
See Conclusions or Pending Issues and List of Inadequacies:
Recommendations: No ethical obstacles
Conclusions or Pending Issues and List of Inadequacies:
Submit partial and final research reports according to the CNS/CONEP recommendations;
Develop strategies for disseminating research results to the HGRS scientific community.
Final Considerations at the discretion of the CEP:
SALVADOR, November 21, 2018
Jorge Luis Motta dos Anjos
(Coordinator)
Signed by:
This opinion was prepared based on the documents listed below:
Type Document File Post Author Status
Basic Project Information
PB_BASIC_INFORMATION_OF_PROJECT_847943.pdf
Accepted
Opinion Status:
Approved
Requires CONEP Review:
No

https://plataformabrasil.saude.gov.br/login.jsf

## ACKNOWLEDGMENTS

The authors thank the hyperbaric medicine team and vascular laboratory staff for their dedicated support throughout this study.

